# Disentangling Inequalities in Tobacco Use: A Decomposition of Rural–Urban Differences Among Men in Nigeria

**DOI:** 10.1101/2025.06.29.25330496

**Authors:** Adams Victor Eseoghen, Obinna Princewill Anyatonwu, Jimoh Boluwatife John, Osham Joan Adams

## Abstract

**Introduction:** Tobacco smoking is a leading contributor to preventable mortality globally, with deaths projected to reach 10 million annually by 2030. While historically more prevalent in high-income countries, recent evidence indicates a rising burden of tobacco use in low– and middle-income countries, including Nigeria. This study aimed to examine rural-urban disparities in tobacco use among Nigerian men and identify the key contributing factors.

**Methods:** This cross-sectional study utilized data from the most recent Nigeria Demographic and Health Survey (NDHS), comprising 13,311 men. We carried out descriptive statistics and the Blinder-Oaxaca decomposition technique to assess differences in tobacco use between rural and urban populations and to quantify the contribution of materialistic, behavioral/cultural, and psycho-social factors to the observed disparity.

**Results:** A small but significant 0.4 percentage point difference in tobacco use was observed between rural and urban men. The materialistic perspective, primarily household wealth and educational attainment, explained the majority of the disparity. Men from the richest and richer households contributed 417.3% and 129.8% to the gap, respectively, while primary and secondary education accounted for 34.4% and 143.2%. Contrary to prior studies, urban men with higher education levels reported higher tobacco use than their rural counterparts. Behavioral/cultural and psycho-social factors, including religion and marital status, helped reduce the rural-urban tobacco use gap. Specifically, being Muslim was associated with lower tobacco use, likely due to cultural and religious norms.

**Conclusion:** Although the rural-urban disparity in tobacco use among Nigerian men is modest, it is primarily driven by materialistic factors such as wealth and education. These findings emphasize the need for targeted policy interventions focusing on socioeconomic inequalities, alongside strengthened enforcement of tobacco control laws, to reduce tobacco use in both rural and urban populations.

**Key Messages:** *What is already known about the topic?:* Tobacco use is a major contributor to preventable death globally, with an increasing burden shifting toward low– and middle-income countries (LMICs) like Nigeria. Prior research indicates that rural populations in LMICs often have higher tobacco use due to factors like poverty, lower education, and limited health information.

*What this study adds:* This study is among the first to use the Blinder-Oaxaca decomposition method to analyze rural–urban differences in tobacco use among Nigerian men. Uniquely, it finds that urban men with higher education levels report higher tobacco use, contradicting global and earlier local trends. Cultural and religious factors (e.g., Islam) and psychosocial elements (e.g., occupation and marital status) reduced the disparity.

*How the study might affect research, practice, or policy:* This study encourages further exploration of why higher-educated urban men are smoking more, suggesting a potential shift in the social meaning or pressures associated with smoking. This calls for tailored tobacco cessation interventions that consider urban wealth and education as emerging risk markers, particularly among men aged 25 – 34 years.

## Introduction

Tobacco smoking is a major contributor to preventable death globally, resulting in about 8 million deaths per year.^1^ Recent evidence suggests that this figure has risen to 8.7 million deaths globally^2^ and are estimated to reach about 10 million annually by 2030.^3^ Tobacco smoking has been linked to a rising burden of lung disease, cardiovascular disease.^1,4^

Historically, tobacco use was most prevalent in high-income Western European nations, with rates as high as 37% among males and 25% among females.^5^ However, studies have shown that low– and middle-income countries (LMICs) are now experiencing a surge in tobacco use. Currently, about 80% of the world’s tobacco users reside in LMICs^6^, a trend that is contributing substantially to rising tobacco-related morbidity and mortality in these regions.^7^ This observed shift in tobacco use is even more prominent in Sub-Saharan Africa, mainly due to the affordability of cigarettes and aggressive marketing by tobacco companies, socio-cultural dynamics, lack of awareness and poor implementation of existing regulations and tobacco laws.^8^

Nigeria, a middle-income country in West Africa and one of the continent’s most populous nations, is home to one of Africa’s largest tobacco markets, with over 18 billion cigarettes sold annually. This widespread availability has contributed to the epidemic spread of tobacco use, particularly among adolescent males^9^, making tobacco a significant cause of preventable illness and death in the country. Although regulatory measures have been introduced to restrict tobacco advertising, packaging, and public smoking,^10^ these efforts have not yielded adequate results.

A 2019 systematic review of 64 studies estimated the unadjusted prevalence of current smokers in Nigeria at 10.4% (9.0%–11.7%), with a markedly higher rate among men (13.4%) compared to women (3.6%).^11^ This gender disparity mirrors global trends, where men consistently account for the majority of tobacco users. Multiple factors influence men’s decision to smoke, ranging from psychological to socio-cultural drivers. A common misconception is that smoking aids in weight control, while others may adopt the habit as a coping mechanism for low self-esteem or as a means to gain social acceptance.^12^ Beyond these individual-level reasons, research has also highlighted a deeper link between smoking and masculinity. Studies suggest that smoking is often used by men to express or reinforce hegemonic masculine traits while simultaneously distancing themselves from behaviours perceived as feminine.^13^

A 2025 study examining regional differences and determinants of tobacco use in Nigeria found that men living in urban settlements had a lower likelihood of using tobacco compared to those in rural areas.^14^ This disparity may be attributed to high poverty levels, greater unemployment, lower education, limited access to health information and specialized care in rural areas, and a stronger presence of tobacco control policies in urban areas.^15^ Consequently, it is not surprising that rural populations exhibit higher rates of health-related risk behaviors compared to urban dwellers.

Additionally, the gradient of stressors across different living environments plays a significant role in smoking behavior. Chronic stressors such as financial difficulties, high-pressure jobs, low perceived control over life, and lack of social support are directly related to smoking and partially mediate the relationship between educational attainment and smoking habits.^16^ In essence, socially disadvantaged individuals, particularly those in rural areas, often endure more stressful conditions, which correlates with higher smoking rates. Hence, in this study, we investigated the factors influencing the rural – urban disparities in tobacco smoking among men in Nigeria.

## Materials and methods

### Study design and data collection

This study is a secondary analysis of cross-sectional data from the 2018 Nigeria Demographic and Health Survey (NDHS), conducted between August and September 2018. The NDHS is nationally representative, sampling approximately 42,000 households across Nigeria.^17^ Using the 2006 Population and Housing Census as its sampling frame, the survey employed a two-stage stratified sampling design across 74 strata (urban and rural areas in each of the 36 states and the Federal Capital Territory). In the first stage, 1,400 enumeration areas were selected with probability proportional to size; in the second, 30 households were randomly selected from each area. A subsample of 13,422 men aged 15–59 was identified, with 13,311 successfully interviewed. Data were collected via computer-assisted personal interviews using standardized questionnaires translated into Igbo, Hausa, and Yoruba and administered by trained personnel.

### Study settings

Nigeria, a lower-middle-income country in Sub-Saharan Africa, is located in the farthest reaches of the Gulf of Guinea on Africa’s west coast. It is bordered by Benin on the West, Cameroon on the East, Chad on the Northeast, and Niger in the far North. Nigeria is one of Africa’s most densely populated countries, with around 212 million people living in an area of 923,768 km2 (356,669 sq. mi). Nigeria is administratively organized into 36 states and the Federal Capital, Abuja. In terms of geopolitical zones, Nigeria is grouped into North-East, North-West, North-Central, South-West, South-East, and South-South.

## Measures

### Outcome Variable

The outcome variable was tobacco use. The variable was derived by asking male respondents if they had used any form of tobacco. Male respondent who answered yes to this question was coded as “1” while those who answered no was recorded as “0”

### Exposure Variable

The primary exposure variable was the place of residence. Participants were categorized into two groups based on their responses: rural or urban residents, irrespective of their geographical region in Nigeria. Those who reported living in rural areas were coded as “1,” while those residing in urban areas were coded as “2.”

### Predictor Variable

The Demographic and Health Survey (DHS) dataset, along with existing literature, guided the selection of predictor variables with an established association with tobacco use.^18^ The final set of variables was determined based on availability in the DHS dataset. Drawing on three theoretical perspectives, the variables were grouped into three categories: materialistic, behavioural/cultural, and psychological. Materialistic variables included educational attainment and household wealth. Education was categorized into four levels: no formal education (reference category), primary, secondary, and higher than secondary. Household wealth was measured using the DHS wealth index, which classified households into five quintiles: poorest (reference), poorer, middle, richer, and richest. Behavioural/cultural variables comprised religion and ethnicity. Religion was grouped into Christianity (reference category), Islam, and no religion. Ethnicity was categorized into ten groups: Fulani (reference), Hausa, Ibibio, Igala, Ijaw, Kanuri, Tiv, Yoruba, and Others.

Psychosocial variables included marital status and occupation. Marital status was divided into never married (reference), married, living with a partner, widowed, divorced, and separated. Occupation was classified into: not working (reference), professional/technical/managerial, clerical, sales, services, skilled manual, unskilled manual, agriculture, and other. Additionally, age was included as a covariate due to its known influence on tobacco use patterns. It was grouped into four categories: 15–24 years (reference), 25–34 years, 35–44 years, and 45–59 years.

## Data analysis

Descriptive statistics were used to summarize tobacco use patterns across rural and urban populations, presented as frequencies and percentages for each predictor variable. To account for potential biases arising from the complex sampling design and challenges during data collection, sample weights provided in the DHS dataset were applied. This study utilized the Blinder-Oaxaca decomposition method to examine the rural–urban differences ^19, 20^ in tobacco use among men in Nigeria. The Blinder-Oaxaca decomposition method was first implemented on the Stata software by Jann.^21^ This statistical technique separates the overall difference in smoking prevalence between the two groups into two components: (1) the portion explained by differences in the distribution of predictor variables (e.g., education, wealth), and (2) the portion due to differences in the effects of these variables on smoking behaviour. The analysis followed a two-fold decomposition process: first, estimating the overall rural–urban gap in tobacco use; and second, quantifying the contribution of each explanatory variable to the observed disparity using coefficients from a pooled regression model. To check for multicollinearity, the variance inflation factor (VIF) was performed. The average VIF was 1.50, which is well within acceptable limits, implying that multicollinearity in the models is unlikely to be a problem. There were no missing values in any of the variables because they were 100% complete. All analyses were performed using Stata/MP version 13.0.

## Ethical consideration

The survey procedure, protocol and instruments used in the 2018 Nigeria Demographic Health Survey received ethical approval from the Federal Ministry of Health of Nigeria’s National Health Research Ethics Committee and the Ethics Committee of Opinion Research Corporation Macro International, Inc. (ORC Macro Inc., Calverton, MD, USA). Before presenting the questionnaire to the participants, the trained personnel obtained informed consent. During the release of the survey results, any information that could be used to identify the study participants were made anonymous. Permission to use and access the Demographic Health Survey dataset was obtained via written consent of the DHS Program. We obtained permission to use the Demographic Health Survey dataset through online registration and formal application to the DHS Program.

## Results

### Descriptive Statistics

A total of 13,311 males were interviewed, forming the basis of the current study. Table 1 highlights the study participants’ characteristics by place of residence, 7,805 (58.6%) resided in the rural settlement, while 5,506 (41.4%) resided in the urban settlement. Majority of our participant were within the age group 15-24 years (30.2%), whilst the least (19.8%) were within the age 45-59 years.

**Table 1:** Weighted frequencies of predictor variables by place of residence.

| Variables | Rural | Urban | Total |
| --- | --- | --- | --- |
| **Age (years)** |  |  |  |
| 15-24 | 2,438 (31.2%) | 1,581 (28.7%) | 4,019 (30.2%) |
| 25-34 | 2,027 (25.9%) | 1,342 (24.4%) | 3,369 (25.3%) |
| 35-44 | 1,808 (23.1%) | 1,480 (26.9%) | 3,288 (24.7%) |
| 45-59 | 1,532 (19.6%) | 1,103 (20.0%) | 2,635 (19.8%) |
| **Materialistic perspective variables** |  |  |  |
| **Education level** |  |  |  |
| No formal education | 2,455 (31.5%) | 491 (8.9%) | 2,946 (22.1%) |
| Primary | 1,240 (15.9%) | 674 (12.2%) | 1,914 (14.3%) |
| Secondary | 3,243 (41.6%) | 2,957 (53.7%) | 6,200 (46.6%) |
| Higher Education | 867 (11.1%) | 1,384 (25.1%) | 2,251 (16.9%) |
| **Wealth quintiles** |  |  |  |
| Poorest | 2,235 (28.6%) | 196 (3.6%) | 2,431 (18.3%) |
| Poorer | 2,012 (25.8%) | 431 (7.8%) | 2,443 (18.4%) |
| Middle | 1,766 (22.6%) | 1,092 (19.8%) | 2,858 (21.5%) |
| Richer | 1,168 (15.0%) | 1,717 (31.2%) | 2,885 (21.7%) |
| Richest | 624 (8.0%) | 2,070 (37.6%) | 2,694 (20.2%) |
| **Behavioral/Cultural perspective variables** |  |  |  |
| **Religion** |  |  |  |
| Christian | 3,554 (45.5%) | 2,981 (54.1%) | 6,535 (49.1%) |
| Islam | 4,183 (53.6%) | 2,481 (45.1%) | 6,664 (50.1%) |
| Other | 68 (0.9%) | 44 (0.8%) | 112 (0.8%) |
| **Ethnicity** |  |  |  |
| Fulani | 629 (8.1%) | 191 (3.5%) | 820 (6.2%) |
| Hausa | 2,326 (29.9%) | 1,252 (22.7%) | 3,577 (26.9%) |
| Ibibio | 192 (2.5%) | 59 (1.1%) | 250 (1.9%) |
| Igala | 101 (1.3%) | 66 (1.2%) | 167 (1.3%) |
| Igbo | 770 (9.9%) | 1,364 (24.8%) | 2,134 (16.0%) |
| Ijaw | 236 (3.0%) | 114 (2.1%) | 350 (2.6%) |
| Kanuri | 203 (2.6%) | 96 (1.7%) | 299 (2.3%) |
| Tiv | 215 (2.8%) | 35 (0.6%) | 250 (1.9%) |
| Yoruba | 510 (6.5%) | 1,301 (23.6%) | 1,811 (13.6%) |
| Others | 2,623 (33.6%) | 1,029 (18.7%) | 3,652 (27.4%) |
| **Psychosocial perspective variables** |  |  |  |
| **Marital Status** |  |  |  |
| Never married | 2,883 (36.9%) | 2,222 (40.4%) | 5,105 (38.4%) |
| Married | 4,697 (60.2%) | 3,041 (55.2%) | 7,738 (58.1%) |
| Living with partner | 96 (1.2%) | 184 (3.3%) | 280 (2.1%) |
| Widowed | 34 (0.4%) | 22 (0.4%) | 56 (0.4%) |
| Divorced | 51 (0.7%) | 18 (0.3%) | 69 (0.5%) |
| Not living together | 44 (0.6%) | 19 (0.4%) | 63 (0.5%) |

| Occupation Type |  |  |  |
| --- | --- | --- | --- |
| Not working | 724 (9.3%) | 741 (13.5%) | 1,465 (11.0%) |
| Professional | 530 (6.8%) | 907 (16.5%) | 1,437 (10.8%) |
| Clerical | 89 (1.1%) | 157 (2.9%) | 246 (1.9%) |
| Sales | 770 (9.9%) | 926 (16.8%) | 1,696 (12.7%) |
| Services | 483 (6.2%) | 546 (9.9%) | 1,029 (7.7%) |
| Skilled manual | 538 (6.2%) | 740 (13.4%) | 1,278 (9.6%) |
| Unskilled manual | 341 (4.4%) | 378 (6.9%) | 719 (5.4%) |
| Agriculture | 4,308 (55.2%) | 1,082 (19.7%) | 5,390 (40.5%) |
| Others | 22 (0.3%) | 29 (0.5%) | 51 (0.4%) |

For the materialistic perspective variables, most participants had at least a secondary level of education (46.6%), with 41.6% residing in rural area while 53.7% lived in urban areas. A smaller proportion had only primary education, with 15.9% residing in rural areas and 12.2% in urban areas. Notably, a large proportion of rural men (28.6%) belonged to the poorest households, compared to just 3.6% in urban settings.

In terms of behavioral and cultural characteristics, most respondents were Muslim (50.1%), with a higher proportion living in rural areas (53.6%) compared to urban (45.1%). Additionally, 27.4% identified with other ethnic groups, predominantly from rural areas (33.6%) versus 18.7% in urban locations Regarding psychosocial variables, the majority of respondents were married, with 60.2% residing in rural areas and 55.2% in urban areas. Most respondents were also employed in the agricultural sector, particularly in rural areas (55.2%) compared to urban areas (19.7%).

### Blinder-Oaxaca Decomposition Analysis

The results of the explanatory variables, as well as the decomposition results of individual and grouped variables, are presented in Table 2. Overall, the rural-urban differences in tobacco use among men accounted for 0.4 percentage points. However, this difference was not significant. The rural-urban disparity in tobacco usage among men in Nigeria was explained by 1.5% percentage points, whereas –1.2% percent was left unexplained for, which reduces this rural urban disparity.

**Table 2:**
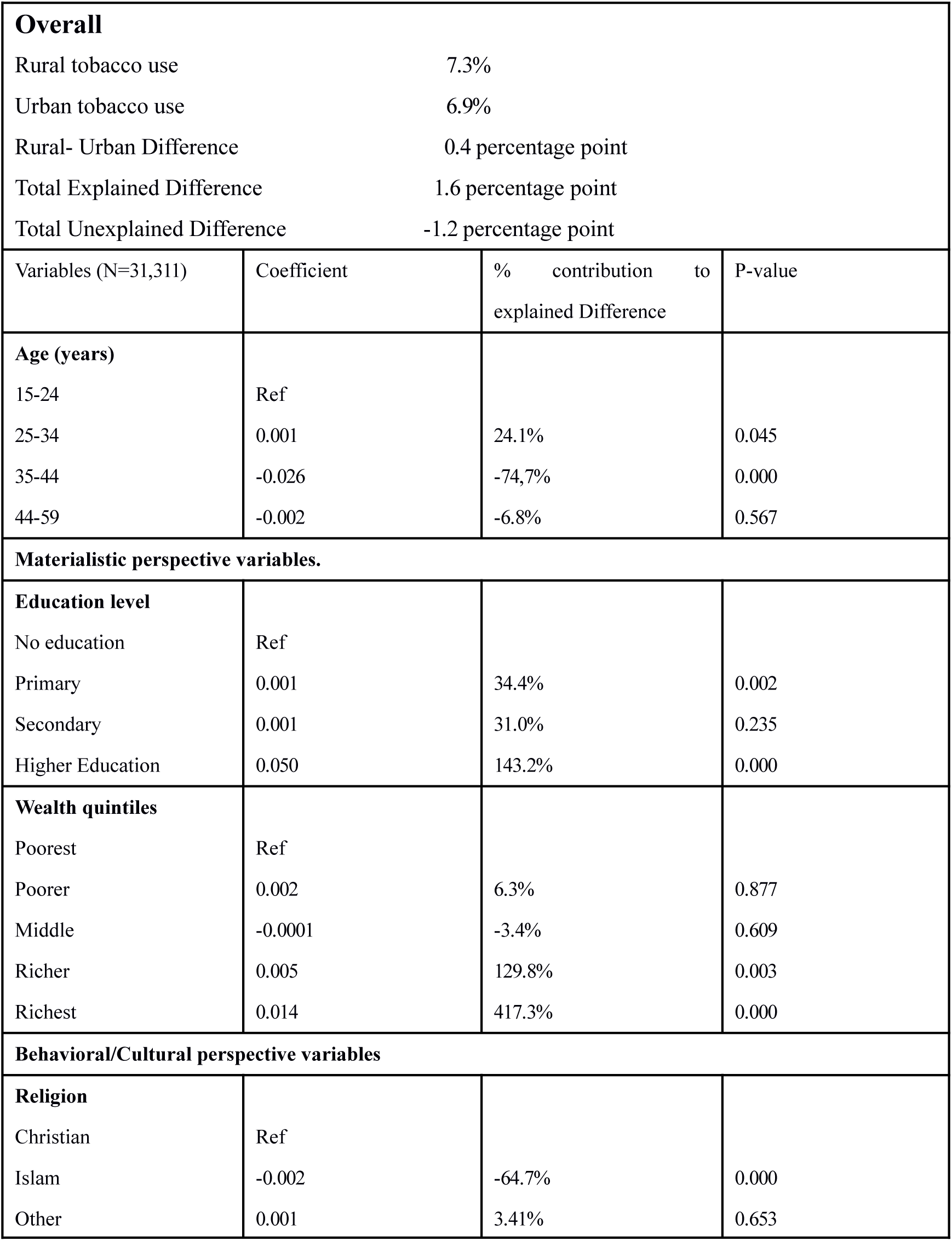
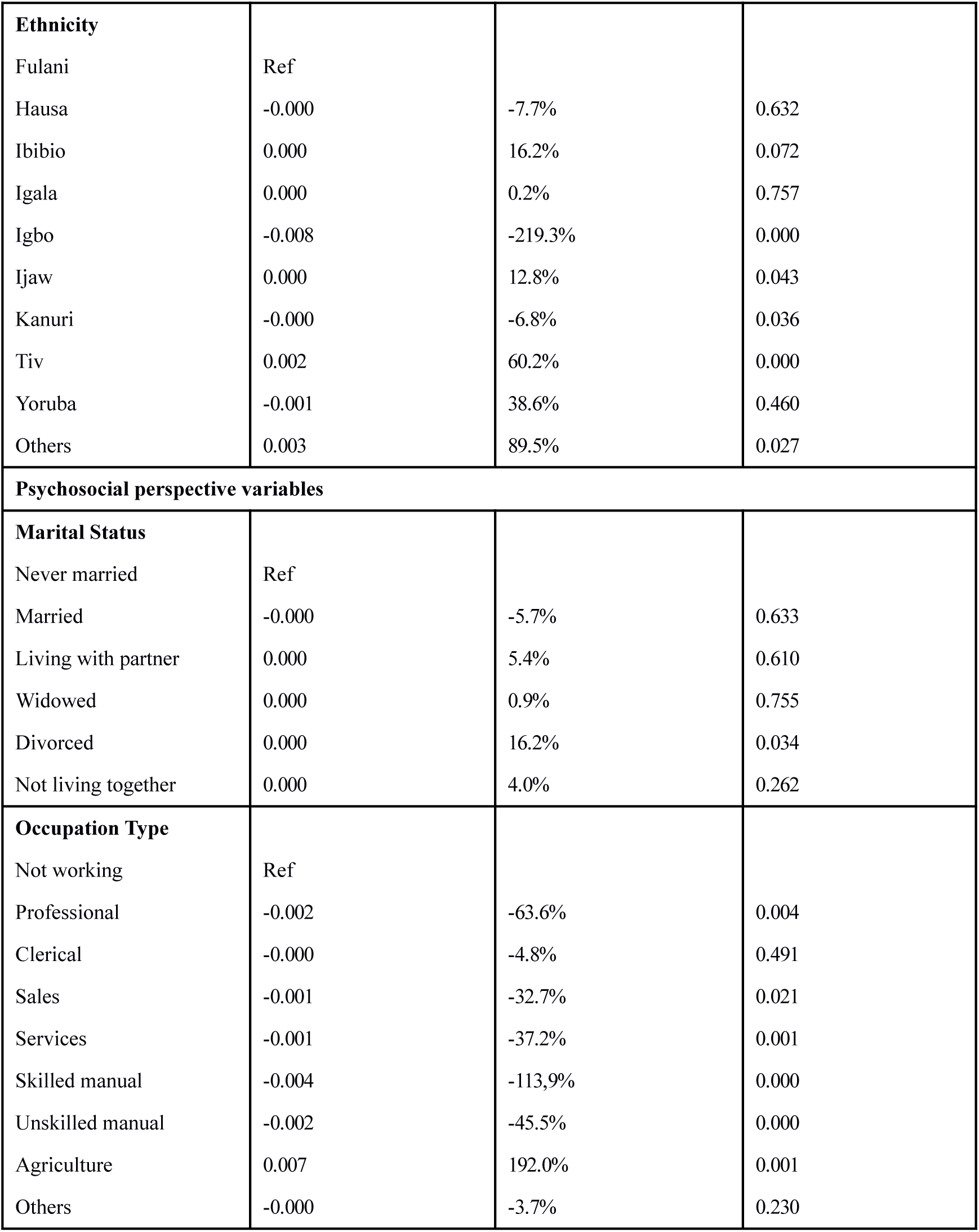
Weighted decomposition of the rural-urban tobacco use disparity among men in Nigeria.

| **Overall** |  |  |  |
| --- | --- | --- | --- |
| Rural tobacco use |  | 7.3% |  |
| Urban tobacco use |  | 6.9% |  |
| Rural- Urban Difference |  | 0.4 percentage point |  |
| Total Explained Difference |  | 1.6 percentage point |  |
| Total Unexplained Difference |  | -1.2 percentage point |  |
| Variables (N=31,311) | Coefficient | % contribution to explained Difference | P-value |
| **Age (years)** |  |  |  |
| 15-24 | Ref |  |  |
| 25-34 | 0.001 | 24.1% | 0.045 |
| 35-44 | -0.026 | -74.7% | 0.000 |
| 44-59 | -0.002 | -6.8% | 0.567 |
| **Materialistic perspective variables.** |  |  |  |
| **Education level** |  |  |  |
| No education | Ref |  |  |
| Primary | 0.001 | 34.4% | 0.002 |
| Secondary | 0.001 | 31.0% | 0.235 |
| Higher Education | 0.050 | 143.2% | 0.000 |
| **Wealth quintiles** |  |  |  |
| Poorest | Ref |  |  |
| Poorer | 0.002 | 6.3% | 0.877 |
| Middle | -0.0001 | -3.4% | 0.609 |
| Richer | 0.005 | 129.8% | 0.003 |
| Richest | 0.014 | 417.3% | 0.000 |
| **Behavioral/Cultural perspective variables** |  |  |  |
| **Religion** |  |  |  |
| Christian | Ref |  |  |
| Islam | -0.002 | -64.7% | 0.000 |
| Other | 0.001 | 3.41% | 0.653 |
| **Ethnicity** |  |  |  |
| Fulani | Ref |  |  |
| Hausa | -0.000 | -7.7% | 0.632 |
| Ibibio | 0.000 | 16.2% | 0.072 |
| Igala | 0.000 | 0.2% | 0.757 |
| Igbo | -0.008 | -219.3% | 0.000 |
| Ijaw | 0.000 | 12.8% | 0.043 |
| Kanuri | -0.000 | -6.8% | 0.036 |
| Tiv | 0.002 | 60.2% | 0.000 |
| Yoruba | -0.001 | 38.6% | 0.460 |
| Others | 0.003 | 89.5% | 0.027 |
| **Psychosocial perspective variables** |  |  |  |
| **Marital Status** |  |  |  |
| Never married | Ref |  |  |
| Married | -0.000 | -5.7% | 0.633 |
| Living with partner | 0.000 | 5.4% | 0.610 |
| Widowed | 0.000 | 0.9% | 0.755 |
| Divorced | 0.000 | 16.2% | 0.034 |
| Not living together | 0.000 | 4.0% | 0.262 |
| **Occupation Type** |  |  |  |
| Not working | Ref |  |  |
| Professional | -0.002 | -63.6% | 0.004 |
| Clerical | -0.000 | -4.8% | 0.491 |
| Sales | -0.001 | -32.7% | 0.021 |
| Services | -0.001 | -37.2% | 0.001 |
| Skilled manual | -0.004 | -113.9% | 0.000 |
| Unskilled manual | -0.002 | -45.5% | 0.000 |
| Agriculture | 0.007 | 192.0% | 0.001 |
| Others | -0.000 | -3.7% | 0.230 |

Men aged 25–34 and 35–44 played significant roles in explaining the rural–urban disparity in tobacco use among men. Compared to those aged 15–24, men in the 25–34 age group accounted for a 24.1% contribution to the disparity, while those aged 35–44 reduced the disparity by 74.7%.

Taking the materialistic perspective into account, men with primary education (34.4%) and higher education (143.2%) increased the urban-rural disparity in tobacco usage among men when compared to men with no formal education. Furthermore, obtaining a secondary school education also widens the differences between rural and urban men tobacco usage by 30.95 %, however this is not statistically significant.

In terms of wealth, tobacco use was higher among men living in urban areas compared to those in rural areas. Men from the richest (417.3%) and richer (129.8%) households contributed substantially to the rural–urban disparity in tobacco use, with both effects being statistically significant. This suggests that the disparity would be reduced if men in rural areas had similar wealth status to their urban counterparts.

When examining behavioral and cultural factors, Islam was found to significantly reduce the rural–urban disparity in tobacco use among men by 64.7% compared to Christianity. Ethnic differences also played a key role in explaining this disparity. Relative to the Fulani ethnic group, the Ijaw (12.8%), Tiv (60.2%), and ‘Other’ ethnic groups (230.1%) contributed to widening the gap, while the Igbo (−219.3%) and Kanuri (−6.8%) ethnic groups contributed to reducing it.

For the Psychosocial perspective, marital status did not significantly contribute to explaining the rural urban difference of tobacco use among men. Conclusively, working in the Agricultural sector significantly increase the rural urban disparity, the other job type all lowered the rural and urban disparity by compared to not working.

As can be seen from the overall contribution of each theoretical perspective, materialistic factors accounted for a substantial 758.6% of the observed disparity in tobacco use. In contrast, behavioral/cultural factors reduced the disparity by 77.6%, age by 57.4%, and psychosocial factors by 88.6%. (Figure 2)

**Figure 2:**
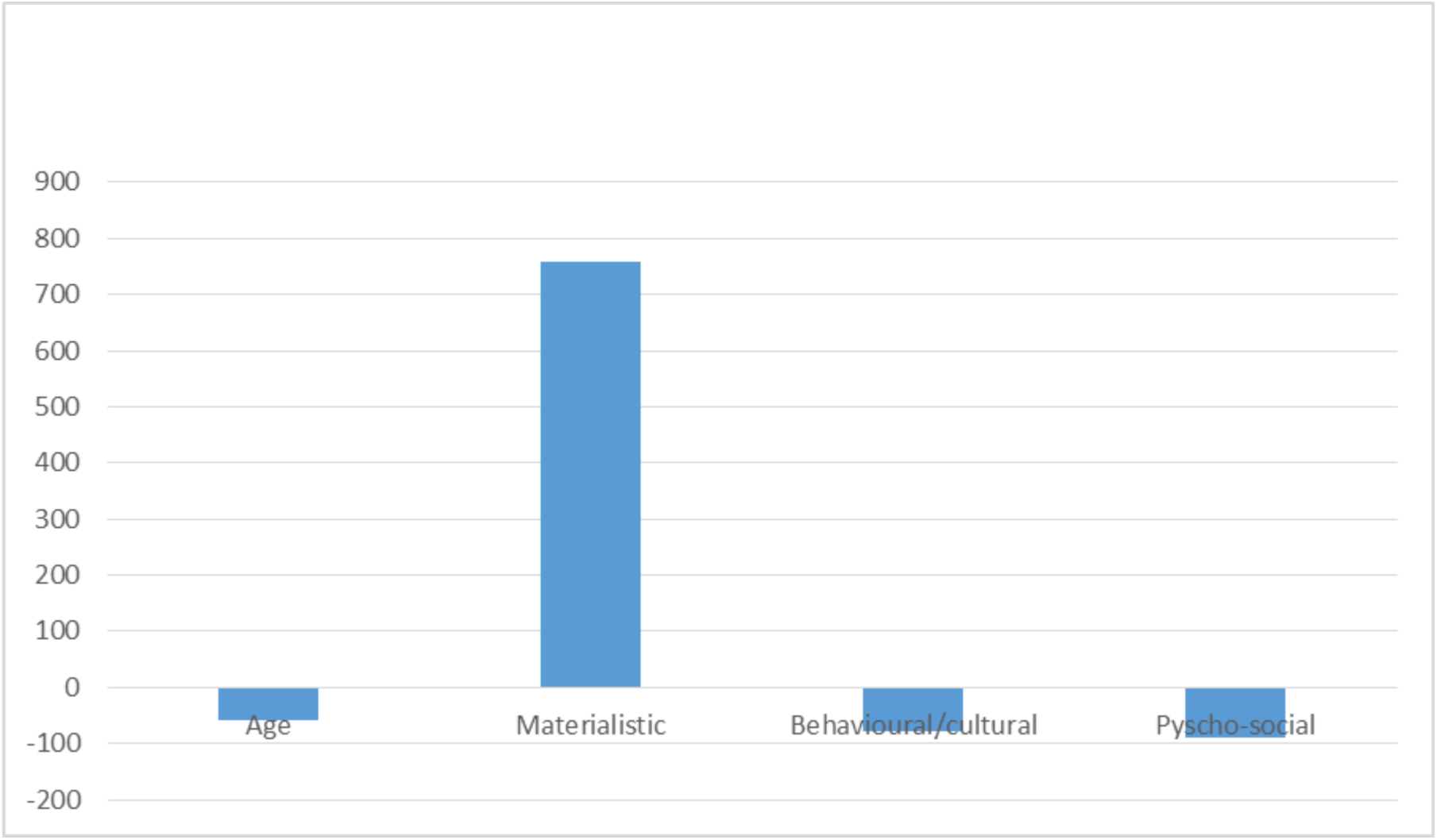
Contribution of age, materialistic, behavioral/cultural, and psycho-social aggregated factors to urban and rural difference.

## Discussion

This present study aimed to examine rural-urban differences in tobacco use among Nigerian men using data from the Nigeria Demographic and Health Survey (NDHS), and to identify the key factors contributing to the observed gap. The analysis showed a 0.4 percentage point difference in tobacco use between rural and urban residents. Materialistic factors, particularly education level and wealth status, accounted for the largest share of this disparity. In contrast, behavioral/cultural and psychosocial factors, such as marital status and specific ethnic group affiliation, narrowed the gap in rural-urban tobacco use.

Participant age generally reduced the disparity in tobacco use between rural and urban men. However, urban men aged 25–34 years still contributed disproportionately to overall tobacco consumption. Several factors may explain this pattern. Firstly, urban environments host a far greater density of kiosks, shops, and street vendors selling cigarettes and other tobacco products, making purchases more convenient than in rural areas. Secondly, tobacco companies concentrate their advertising and promotional efforts in cities, where young adults comprise a key demographic. Thirdly, men aged 25–34 years in urban centres enjoy higher incomes yet face intense work pressures and tight deadlines. Hence, they are more likely to adopt smoking as a stress-coping strategy. In accordance with this present study, Aniwada et al. found that adults in Nigeria aged 25 – 34 years and ≥ 35 years are significantly more likely to use tobacco than their 15–24-year-old counterparts.^22^ This association between age and tobacco use aligns with other related studies. ^23,24^

Wealth also emerged as a driving factor for tobacco use in urban settings. Men in the richest and richer wealth quintiles, compared to their rural counterparts, contributed substantially to this disparity. This means that higher socioeconomic status translates into greater disposable income, making tobacco products readily affordable. This observation contradicts global patterns where higher socioeconomic status is often linked with lower tobacco use. Supporting this, a recent multicountry analysis that examined the socioeconomic inequalities in lifestyle risk factors found that, uniquely in Nigeria, tobacco use is concentrated among the better off whereas in most other LMIC it is pro-poor. ^25^ In Kenya, daily tobacco use disproportionately affects men from poorer urban households compared to their wealthier peers.^26^ By contrast, a study in Vietnam on the socioeconomic inequalities in smoking using decomposition analysis, showed that poorer men are more likely to smoke and smoke more than the better off.^27^

An interesting finding from this study is that educational attainment contributed to widening the gap in tobacco use between rural and urban men. Contrary to expectations and previous Nigerian studies, urban men with higher education levels were found to use more tobacco products than their rural counterparts. Earlier studies have consistently shown that individuals with education beyond secondary school are generally less likely to smoke compared to those with only primary or no education ^22, 28^ as higher education is typically associated with greater awareness of health risks.

This unexpected trend suggests a shift in behavior among educated urban men and highlights the need for further research to understand the social or contextual factors driving tobacco use in this group.

The behavioral/cultural and psycho-social factors were found to significantly reduce the rural-urban tobacco use gap among men. Although religiosity regardless of whether affiliation increases the likelihood of abstaining from tobacco use^29^, the observed difference can be attributed to stricter religious doctrines and community norms more commonly enforced within the Islamic communities. In many Islamic communities, tobacco consumption is not only discouraged but often explicitly prohibited, as it is perceived to harm the body. Additionally, Islamic communities often have stronger social control mechanisms, where adherence to religious norms is closely monitored and deviations are discouraged.^30^

Our findings further revealed that being divorced widened the rural – urban gap in tobacco use, while being married contributed to narrowing it. This trend is not surprising, as being divorced regardless of setting is often accompanied by emotional distress, social isolation, and financial strain.^31,32^ In urban, fast-paced environments where life pressures such as demanding work conditions and high living costs are more intense, tobacco use may serve as a coping mechanism^14^ for divorced men. The emotional toll of separation, compounded by the stressors of city life, likely contributes to higher tobacco use. This finding aligns with existing research on the relationship between marital status and tobacco use, which depicts that adults who are separated or divorced are nearly twice as likely to smoke compared to those who are still married.^33^

In light of these findings, and to accelerate progress toward the tobacco endgame, the Nigerian government should fully implement the WHO MPOWER demand reduction package, which includes increasing tobacco taxes, enforcement of smoke-free regulations, prohibiting all advertising, promotion, and sponsorship, mandating plain packaging and graphic warning labels, funding public education, and expanding cessation services. Furthermore, it must uphold larger FCTC obligations by protecting health policies from industry manipulation, strengthening tobacco control governance, countering illegal trafficking, regulating product contents, and implementing age-of-sale requirements.^34^ The evidence of this aforementioned policy recommendation has been noticed in related studies.^35,36^

Our study has its strength and limitations. The study’s key strength lies in its use of nationally representative Demographic and Health Survey (DHS) data, covering all 36 states in Nigeria, with no missing values across variables organized under material, cultural/behavioral, and psychosocial frameworks. Given the stigma connected with tobacco smoking, the issue of tobacco use in Nigeria is extremely delicate, as tobacco use was self-reported, the use, frequency, and type of tobacco use in Nigeria may be prone to response bias and underreporting. Furthermore, the cross-sectional design also limits the ability to infer causality.

## Conclusion

This study reveals modest rural – urban differences in tobacco use among Nigerian men, primarily driven by material factors. Targeted interventions are needed, particularly for urban men aged 25–34 years and those in higher socioeconomic groups who represent emerging risk patterns. Policymakers must adopt a multidimensional approach that considers not only individual behaviours but also the broader social, cultural, and economic contexts. Integrating regulatory measures with culturally tailored public health strategies such as strengthening education, addressing urban lifestyle pressures, and leveraging religious and community norms will be key in making progress. Continued research and surveillance are essential to track evolving trends and inform responsive tobacco control policies.

## Data Availability

All data produced in the present work are contained in the manuscript

